# Changes in work and health of Australians during the COVID-19 pandemic: a longitudinal cohort study

**DOI:** 10.1101/2021.05.02.21256492

**Authors:** Daniel Griffiths, Luke Sheehan, Caryn van Vreden, Dennis Petrie, Peter Whiteford, Malcolm R Sim, Alex Collie

**Affiliations:** School of Public Health and Preventive Medicine, Monash University, Australia; Centre for Health Economics, Monash University, Australia; Crawford School of Public Policy, Australian National University, Australia

**Keywords:** COVID-19, mental health, unemployment, physical health

## Abstract

**Objectives:** To determine the long-term effects of work loss on health during the COVID-19 pandemic, and whether any effects are persistent upon returning to work.

**Methods:** A prospective longitudinal cohort study of 2603 participants across Australia monitored changes in health and work during between March and December 2020, with participants completing surveys at baseline and 1, 3 and 6 months later. Outcomes described psychological distress, and mental and physical health. Linear mixed regression models examined associations between changes in health and experiences of work loss, and return to work, over time.

**Results:** Losing work during the early stages of the pandemic was associated with long-term poorer mental health, which began to recover over time as some returned to work. Physical health deteriorated over time, greater for people not working at baseline. Being out of work was associated with poorer mental health, but better physical health. These effects were larger for people that had recently lost work than for people with sustained work loss, and retaining employment played a protective role. Generally, returning to work resulted in poorer physical health and improvements in mental health, although this depended on the broader context of changes in work.

**Conclusions:** Work cessation during the pandemic led to poor health outcomes and had long-lasting effects. Returning to work benefits mental health but may reduce physical activity in the short-term. We encourage the provision of accessible mental health supports and services immediately following loss of work, and for people with prolonged forms of work loss.

## Introduction

Involuntary job loss can be a devastating experience, affecting livelihoods, mental health and a sense of identity. Wide-scale job loss has been a feature of the COVID-19 pandemic, arising from public health measures designed to contain viral spread such as business closures and movement restrictions and physical distancing. These restrictions are now easing in some countries as rates of vaccination increase, however the full recovery of lost working hours is expected to take several years, with a projected recovery in 2021 still resulting in 36 to 130 million full-time equivalent job losses compared to pre-pandemic levels [1]. As people return to work, we can begin to determine whether some of the immediate health consequences of being out of work during the early pandemic, such as worsened mental health [2], either quickly recalibrate upon returning to work, or persist for an extended period of time. Australia is a unique setting for such a study, having experienced an initial national lockdown resulting in widescale loss of working hours, but with business conditions improving rapidly late in 2020 thanks to a largely successful suppression of COVID-19 community transmission.

Within Australia, as in other nations, the recovery of jobs and working hours are not evenly distributed across the labour force. Some people are able to quickly return to work as restrictions ease, whilst others may continue to struggle to find employment for prolonged periods. Following the first national wave of COVID-19 in Australia, casual workers (i.e. employees without sick or holiday leave entitlements) accounted for around two-thirds of people losing their job, and casual employment recovered by 37.2% between May and August 2020 [3], coinciding with easing restrictions across most of the country. However, from July to October 2020, the state of Victoria experienced an extended community lockdown during a second wave of COVID-19 localised within the state. Job losses were also more pronounced in Victoria [4]. The health impacts of Victorian workers are likely to differ due to the experience of an extended and stringent lockdown, in addition to a larger proportion of individuals experiencing multiple periods of work cessation and return to work. Additionally, some will have sustained longer-term unemployment or loss of work throughout most of the year, and the health outcomes of this group may differ. Nationally, some businesses that survived the initial national lockdowns in March/April were not able to sustain operations and closed later in the year, while others re-opened in a very different operating environment and in a reduced capacity. In summary, many workers have moved in and out of work during the pandemic, following the initial nation-wide work loss observed in March / April 2020.

The increased burden of mental ill health has been a feature of the pandemic [5], due in part to high levels of social isolation, restrictions on everyday activities, and the loss of work. Returning to work is generally positive for mental health [6], although this is not well described in a pandemic context. There is also limited understanding of how mental health is affected by multiple instances of work loss. for instance in response to repeated business closure. The negative long-term impacts of unemployment on mental health have been characterised outside of the COVID-19 context, including studies of multiple exposures to unemployment [7], and others observing how mental health itself influences longer durations of unemployment [8]. The unique circumstances of the pandemic may alter these relationships. During the pandemic, people are not only experiencing the loss of work, but are also facing uncertainty about returning to work amidst a COVID-induced recession along with regular and at times sudden changes in public health preventive measures that may affect work opportunities. Employees that are stood-down from work and supported by temporary wage subsidies [9], intended to counteract labour market effects, may have increased confidence in returning to work as the economy recovers and as restrictions are eased, which may have secondary implications for health.

The pandemic has also had a marked effect on our physical behaviour. The closure of non-essential businesses and restrictions on gatherings have reduced the opportunity for some physical activities like group sports and use of gyms and indoor venues. However, a consequence of such restrictions is the increased availability of discretionary time, particularly for individuals that are out of work. Some jurisdictional lockdown rules and messaging have include time-limitations for exercise, which may promote increased physical activity in some cases [10]. Lockdown rules have also included working from home directives that risks a reduction in incidental exercise associated with travelling to and from a workplace. To our knowledge the long-term impacts of pandemic-related work loss and return to work on physical health have not yet been examined.

We describe a cohort of working-age Australians, focusing on those experiencing work loss, to evaluate the health responses during and following periods of work cessation and upon returning to work throughout the pandemic. Specifically, we sought to answer four research questions:

1. Does being out of work early in the pandemic affect health six months later?

2. Do health impacts differ for people employed but not currently working?

3. What are the health impacts of changes in exposure to work?

4. How does the longitudinal context of changes in work effect health?

## Methods

### Data collection, setting and participants

We report findings from a longitudinal cohort study of people that were employed in a paid job or self-employed prior to the pandemic, residing in Australia and aged at least 18 years.

A total of 2603 participants enrolled in the study and completed a 20-minute baseline survey (either online or via a telephone survey) between 27 March and 12 June 2020, of which 2151 participants also consented to future follow-up surveys at 1, 3 and 6 months after baseline. Baseline survey measures for the cohort have been previously described [2]. The cohort includes a group of people experiencing work loss early in the pandemic and a control group of people who did not lose working hours.

### Health outcomes

Three health outcomes were assessed at each of the four-survey time-points: (1) psychological distress, (2) mental health, and (3) physical health. Psychological distress was assessed using the total scores from the 6-item Kessler Psychological Distress scale [11] ranging from 6 to 30. Mental and physical health was assessed using the mental health component summary score and the physical health component summary score from the 12-item Short Form Health Survey (SF-12) [12], ranging from 0 to 100.

### Work

Exposure to work was dichotomised into two groups at each survey timepoint, where individuals were described as either *Working* (W) or *Not working* (N). The state of *Working* describes people that were employed at the time, and worked more than zero hours during the prior week. The state of *Not working* describes people that had either lost their job and were unemployed, or that had been stood down from work or furloughed.

### Employment

An employment variable was defined to describe whether people were employed or not at each survey time-point. Workers were assigned at each survey time-point as either being employed and working, employed and not working, or unemployed and not working.

### Analytical approach

A total of 6859 observations were available for statistical analysis from 2603 participants across four surveys.

Firstly, summary statistics were calculated to describe subgroups of people that were either working or not working at baseline. Groups were summarised according to demographics, pre-existing health conditions, residential location, and survey mode.

Secondly, linear mixed models were used to account for repeated measures. Four models labelled 1-4 were designed to evaluate health outcomes with different exposures describing work loss, and were designed to answer the four research questions. Model 1 focuses on baseline work status groups and their changes in health over time. Models 2-4 describe health outcomes along with their corresponding work status at any survey time-point. The four models were estimated for each of the three health outcomes.

### Model 1: Does being out of work early in the pandemic affect health six months later?

Model 1 describes health outcomes across each of the four survey time-points, comparing individuals that were either working or not working at baseline. The exposure group for model 1 was working status at baseline (i.e. time-invariant work status) with an interaction term describing each survey time-point allowing us to estimate the differences in health outcomes for both groups at each survey time-point regardless of their future working status.

### Model 2: Do health impacts differ for people not working if they are employed?

Model 2 is separated into two sub-models. Model 2.1 describes the health outcomes for those individuals either working, or not working at each of the four respective survey-time points. The exposure for Model 2.1 was working status at the same survey time-point as the corresponding health score. Model 2.2 is the same as Model 2.1 but with those who were not currently working separated into those who were still employed and those who were unemployed.

### Model 3: What are the health impacts of changes in exposure to work?

The exposure for Model 3 was defined as an interaction between the work status at any given survey time-point with the work status of the prior survey. Four outcomes are possible: (1) working at both time-points, (2) not working at both time points; (3) transition from working to not working; (4) transition from not working to working. Participants were assigned as working before baseline.

### Model 4: How does the longitudinal context of changes in work effect health?

The exposure in Model 4 is a three-way interaction term for the work status of a particular survey with the work statuses for the prior two surveys. The reference group for Model 4 describes people that were currently working and that also were working at the two prior survey time-points. Participants were assumed to be working for two pseudo-survey time-points prior to baseline, supported by study participant eligibility criteria for being involved in paid work prior to the pandemic (and specifically during September to December 2019).

### Fixed effects and random effects

Variables describing gender, age group, survey time-point, residential location, and pre-existing health conditions prior to baseline were included in models as fixed effects. Regression models for mental health and psychological distress included fixed effects for pre-existing anxiety and depression. Regression models for physical health included a variable describing the number of pre-existing medical condition categories as none, one, or two or more. An interaction term was included between survey time-point and whether participants resided in Victoria or the Rest of Australia, in addition to fixed effect for survey time-point itself due to known influences of an extended lockdown on health [13]. Previous analyses describe response categories [2] and also identified differences in health outcomes by survey mode, thus survey mode was also included in all models as a fixed effect. Reference groups for fixed effects were male, ages 35-44 years, no pre-existing medical conditions, residing outside of Victoria, and online survey mode. Intercept estimates correspond to these reference groups.

Across all linear mixed models, repeated measurements were incorporated by including random effects for survey time-point with a unique identification number for each participant.

## Results

A summary of the cohort is described in Table 1 showing differences between people that were working at baseline, and people without work. However, many people either subsequently returned to work, lost work, or were in an out of work on multiple occasions. Across four survey time-points, health outcomes were calculated for people in work on 4463 occasions and for people that were not currently working on 2196 occasions. When evaluating transitions between working states, the most common was for people to report working on both occasions (59.8%), followed by movement from working to not working (work loss; 20.4%), not working on both occasions (sustained work loss; 12.1%), and moving from not working to working (return to work, 7.7%). Table 2 presents a summary of findings from regression models.

**Table 1.** Descriptive statistics by working status at baseline.

|  | **Cohort summary statistics.**<br>**N (column %)** |  |
| --- | --- | --- |
| **Working status at baseline** | **No Work**<br>**N=1154** | **Working**<br>**N=1449** |
| **Gender** |  |  |
| Female | 841 (72.9) | 773 (53.3) |
| Male | 303 (26.3) | 672 (46.4) |
| Non-binary/no gender/unspecified | 10 (0.9) | 4 (0.3) |
| **Age Group** |  |  |
| 18 to 24 years | 131 (11.4) | 114 (7.9) |
| 25 to 34 years | 195 (16.9) | 247 (17.0) |
| 35 to 44 years | 189 (16.4) | 295 (20.4) |
| 45 to 54 years | 293 (25.4) | 354 (24.4) |
| 55 to 65 years | 299 (25.9) | 353 (24.4) |
| Over 65 years | 47 (4.1) | 86 (5.9) |
| **Pre-existing mental health conditions** |  |  |
| Anxiety | 313 (27.1) | 150 (10.4) |
| No Anxiety | 841 (72.9) | 1299 (89.6) |
| Depression | 302 (26.2) | 190 (13.1) |
| No Depression | 852 (73.8) | 1259 (86.9) |
| **Number of pre-existing health conditions** |  |  |
| None | 462 (40.0) | 1013 (69.9) |
| One | 331 (28.7) | 266 (18.4) |
| Two or more | 361 (31.3) | 170 (11.7) |
| **Residential Location** |  |  |
| Victoria | 388 (33.6) | 493 (34.0) |
| Rest of Australia | 754 (65.3) | 944 (65.1) |
| Undetermined | 12 (1.0) | 12 (1.0) |
| **Survey mode** |  |  |
| Online | 900 (78.0) | 323 (22.3) |
| Telephone | 254 (22.0) | 1126 (77.7) |

**Table 2.** Changes in health due to loss of work and return to work during the COVID-19 pandemic.

|  | Number of occasions (%) | Adjusted estimates for differences in health score<br>[95% Confidence Interval] |  |  |
| --- | --- | --- | --- | --- |
| | | Psychological distress<br>($-\beta < 0$: worse distress)<sup>‡</sup> | Mental health<br>($\beta < 0$: poorer health) | Physical health<br>($\beta < 0$: poorer health) |
| ***Model 1. Does being out of work early in the pandemic affect health 6 months later?*** | 6859 (100.0) |  |  |  |
| No Work at baseline * baseline | 1154 (16.8) | **-1.23\*\*** [-1.52, -0.93] | **-2.33\*\*** [-3.01, -1.65] | **0.94\*\*** [0.44, 1.44] |
| No Work at baseline * 1 month | 644 (9.4) | **-0.39\*** [-0.74, -0.05] | -0.37 [-1.20, 0.45] | 0.14 [-0.48, 0.75] |
| No Work at baseline * 3 months | 547 (8.0) | -0.33 [-0.73, 0.08] | -0.20 [-1.17, 0.77] | -0.31 [-1.02, 0.41] |
| No Work at baseline * 6 months | 475 (6.9) | -0.22 [-0.73, 0.29] | -0.16 [-1.36, 1.05] | **-1.29\*\*** [-2.17, -0.41] |
| Working at baseline * baseline | 1449 (21.1) | 0.00 (ref.) | 0.00 (ref.) | 0.00 (ref.) |
| Working at baseline * 1 month | 1002 (14.6) | **0.48\*\*** [0.24, 0.71] | **1.56\*\*** [1.00, 2.12] | 0.04 [-0.37, 0.46] |
| Working at baseline * 3 months | 836 (12.2) | **0.81\*\*** [0.54, 1.08] | **2.11\*\*** [1.47, 2.75] | -0.34 [-0.81, 0.13] |
| Working at baseline * 6 months | 752 (11.0) | **1.16\*\*** [0.86, 1.45] | **2.72\*\*** [2.02, 3.42] | -0.50<sup>†</sup> [-1.01, 0.00] |
| ***Model 2.1. Do health impacts differ for people not working?*** | 6859 (100.0) |  |  |  |
| No work (N) | 2196 (32.0) | **-1.13\*\*** [-1.35, -0.91] | **-2.29\*\*** [-2.80, 1.78] | 0.32<sup>†</sup> [-0.06, 0.70] |
| Work (W) | 4663 (68.0) | 0.00 (ref.) | 0.00 (ref.) | 0.00 (ref.) |
| ***Model 2.2. Do health impacts differ for people not working if they are employed?*** | 6859 (100.0) |  |  |  |
| No work (N) * Unemployed | 1163 (17.0) | **-1.65\*\*** [-1.94, -1.37] | **-3.03\*\*** [-3.68, -2.38] | 0.08 [-0.41, 0.57] |
| No work (N) * Employed | 1033 (15.1) | **-0.70\*\*** [-0.96, -0.44] | **-1.65\*\*** [-2.27, -1.04] | **0.52\*** [0.06, 0.98] |
| Working (W) * Employed | 4463 (68.0) | 0.00 (ref.) | 0.00 (ref.) | 0.00 (ref.) |
| ***Model 3. What are the health impacts of changes in exposure to work?*** | 6588 (100.0) |  |  |  |
| No work to No work (N * N) | 796 (12.1) | **-1.19\*\*** [-1.55, -0.83] | **-2.46\*\*** [-3.27, -1.64] | -0.30 [-0.91, 0.30] |
| Working to No work (W * N) | 1346 (20.4) | **-1.07\*\*** [-1.36, -0.78] | **-2.23\*\*** [-2.88, -1.58] | 0.29 [-0.20, 0.77] |
| No work to Working (N * W) | 509 (7.7) | 0.04 [-0.28, 0.36] | -0.03 [-0.79, 0.73] | -0.54<sup>†</sup> [-1.11, 0.02] |
| Working to Working (W * W) | 3936 (59.8) | 0.00 (ref.) | 0.00 (ref.) | 0.00 (ref.) |
| ***Model 4. How does the longitudinal context of changes in work effect health?*** | 6529 (100) |  |  |  |
| N * N * N | 307 (4.7) | **-1.36\*\*** [-1.88, -0.84] | **-2.56\*\*** [-3.74, -1.37] | **-1.17\*\*** [-2.06, -0.28] |
| W * N * N | 226 (3.5) | **-0.89\*\*** [-1.30, -0.47] | **-2.01\*\*** [-2.96, -1.07] | -0.65<sup>†</sup> [-1.36, 0.06] |
| N * W * N | 53 (0.8) | **-0.92\*** [-1.77, -0.07] | -1.77<sup>†</sup> [-3.81, 0.26] | -0.74 [-2.25, 0.77] |
| W * W * N | 216 (3.3) | **-0.98\*\*** [-1.29, -0.66] | **-2.09\*\*** [-2.80, -1.39] | 0.01 [-0.52, 0.55] |
| N * N * W | 486 (7.4) | 0.40 [-0.11, 0.91] | 0.66 [-0.52, 1.85] | **-1.65\*\*** [-2.54, -0.75] |
| W * N * W | 269 (4.1) | 0.03 [-0.41, 0.47] | -0.06 [-1.09, 0.96] | -0.74<sup>†</sup> [-1.51, 0.03] |
| N * W * W | 1291 (19.8) | 0.32 [-0.15, 0.80] | 0.62 [-0.50, 1.73] | **-1.39\*\*** [-2.22, -0.55] |
| W * W * W | 3681 (56.4) | 0.00 (ref.) | 0.00 (ref.) | 0.00 (ref.) |

<sup>†</sup>0.05≤p<0.1, \*0.01≤p<0.05, \*\*p<0.01. <sup>‡</sup>Coefficients and confidence intervals listed for psychological distress within the table have been negated (i.e. -β) to ease comparisons with corresponding coefficients for mental health. Models controlled for gender, age group, pre-existing health conditions, survey time-point, and interactions of survey time-point with residential location (i.e. Victoria or the Rest of Australia); N = No work; W = Working.

### Long term health effects of work loss during the early pandemic

Overall, mental health improved over time and physical health deteriorated (Figure 1, Model 1, Table 2). At baseline mental health scores were lower than pre-pandemic population average levels (i.e. 50/100), whereas physical health scores were notably higher than pre-pandemic population averages throughout all survey time-points.

**Figure 1.**
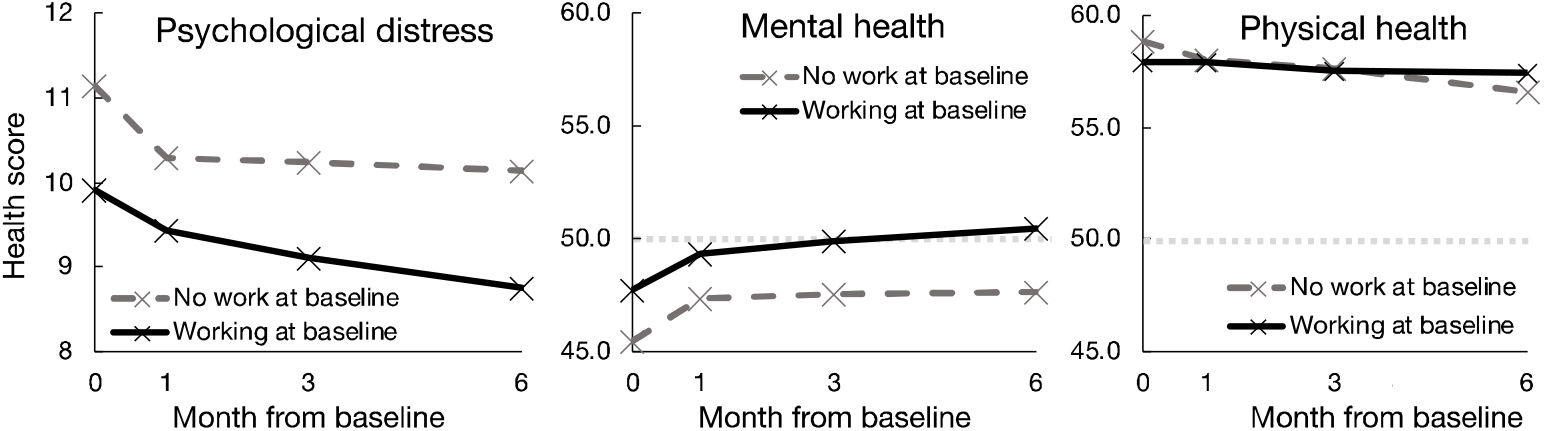
**Adjusted estimates for psychological distress, mental health and physical health scores over 6 months based on baseline work status (i.e. Model 1). Intercept estimates describe covariate reference groups**.

People that were out of work at baseline had higher levels of psychological distress, poorer mental health and better physical health compared to people working at baseline, which continued over 6 months. The reference group of those working at baseline showed poorer physical health after six months compared to their baseline levels. The physical health of those not working at baseline started off better than those working at baseline but ended up worse in comparison after 6 months, following a more rapid deterioration.

### Employment as a protective factor for health during work loss

Throughout the study period, not working was associated with increased psychological distress and poorer mental health compared to being in work (Model 2.1, Table 2). Health outcomes differed between people that were unemployed and people that were employed but not working (Model 2.2, Table 2). Compared to people working, employed individuals who were not working had higher levels of distress, and poorer mental health. These effects were greater among unemployed people. The physical health of employed people who were not working was significantly higher than for people employed and working.

### Persistent health effects following work loss or return to work

The impacts of current working status on health differed depending on prior work status (Model 3, Table 2). Individuals with sustained work loss (i.e., not working at two consecutive time-points) demonstrated the highest levels of distress, and poorest mental health. The negative impacts of not working on mental health and distress were smaller in magnitude for people experiencing work loss (transitioning from working to not working), though still statistically significant.

Findings from Model 4 extend upon Model 3, by showing that the negative effects on mental health and psychological distress are larger when work loss is more acute (i.e. WWN), with smaller effects for sustained work loss (i.e. WNN). The worst mental health outcomes were observed for the continuously out of work group (i.e. NNN), as well as the most elevated levels of psychological distress, and low levels of physical health. Sustaining work (i.e. WWW) was associated with the highest levels of mental and physical health, however those newly returning to work (i.e. NNW) were amongst those with the lowest levels of physical health, which improved with ongoing work (i.e. NWW, and subsequently WWW).

## Discussion

Our findings suggest that the negative mental health consequences of work loss during the COVID-19 pandemic are heightened immediately following loss of work, and that mental health further deteriorates during more prolonged periods of sustained work loss. Whilst previous studies have shown similar patterns comparing people experiencing short-term and long-term unemployment [14], we observe that this extends to include other forms of work cessation like being temporarily stood down from work, and our findings fall within the unprecedented COVID-19 pandemic context. We also describe significant short-term increases in the physical health of those not working but employed, consistent with previous studies on people that are out of work [15]. One hypothesis for this finding is that improved physical health may be a consequence of an increase in time available for physical activity, coupled with an increased perception of job security, and income security for some. We also demonstrate evidence that returning to work is associated with poorer physical health in the short term as people may need to adapt to new jobs, or a change in lifestyle, that may be more physically demanding, which people may become more accustomed to upon continued engagement in work.

Our analyses demonstrate that returning to work during the COVID-19 pandemic is good for mental health and contributes to reduced psychological distress. Returning to work, may reinstate aspects of working that are good for mental health such as greater social interaction, providing a sense of purpose and identity, along with improved financial security. However, we note that returning to workplaces during the pandemic is qualitatively different to a pre-pandemic context, given the large-scale changes to workplace health and safety policies and practices [16] and the backdrop of stress and social disruption. The majority of workers express concerns about returning to workplaces during the pandemic and this may act to reduce some of the mental health benefits that would otherwise be achieved [17]. Continuing to monitor the health of workers once they have returned to work will be essential to understand the longer-term impacts of work loss beyond the initial return to work period.

We have shown evidence that retaining employment when not working potentially moderates the relationship between work loss and mental health, cushioning reductions in mental health by about a half. Whilst unemployed individuals may be eligible to access social security supports, the prospects of securing work during an economic recession will be lower for the unemployed than for people who have retained an employment relationship. We note that furloughed workers may also fear that being stood down represents an interim step on the path to job loss. Consultation and communication between employers and employees about returning to the workplace and future opportunities for re-engagement in work can help to alleviate worker distress in these circumstances [18]. One implication of our findings is that wage subsidy programs and payments that keep people employed whilst businesses close for a temporary period [9] will contribute towards reducing the negative mental health impacts of work loss. It follows that the withdrawal of such economic supports is likely to have negative mental health consequences.

One implication of our findings is that the greatest need for mental health supports and services is in the acute period immediately following loss of work. Provision of such supports has potential to reduce both the short and longer-term mental health consequences of work loss. In Australia, the setting for this study, and in many other nations, there have been additional investment in such services by governments during the pandemic. We previously observed that a relatively small proportion of workers with psychological distress reported accessing formal supports such as psychology/counselling and calling telephone support hotlines [19]. Our findings suggest that the newly unemployed, and people recently stood down or laid off, are a section of the community that will benefit from greater access to, or use of, such services.

To our knowledge, this study reports the first description of longitudinal changes in health and work experienced by Australian workers during the COVID-19 pandemic, which are likely to apply to other countries with similar pandemic-induced changes in work arrangements. One strength of the study is its longitudinal and national coverage of the pre-vaccination phase of the COVID-19 pandemic. This phase consisted of extended and short (or ‘snap’) lockdowns resulting in multiple phases of work loss and work recovery. The longitudinal design enabled us to track the health impacts of moving in and out of work over this time period. We also recruited a comparison group whose working hours were unaffected throughout but also experienced the pandemic environment. Characterising the specifics of work loss can be a challenge, particularly for casual workers with multiple jobs and flexible hours, so our analysis has been limited to describing the impact of working or not working.

Further analysis will be required to understand a more complete range of working circumstances, for example the impacts of partial work loss.

We describe the nature of work loss and return to work within the context of the pandemic environment experienced in Australia, which experienced relatively low rates of infection, and this may differ in countries with larger infection risks, different restrictions and fewer or no social security for individuals out of work. When losing work, we know that social interactions and financial resources can help to minimise some of the risks of poor health outcomes during the early pandemic [2]. The easing of restrictions that enable greater social interaction are anticipated to contribute towards improvements in mental health. However, as we move into the second year of the COVID-19 pandemic, additional factors are likely to shape working lives. Vaccination of the workforce, the withdrawal of some temporary financial supports, and employers approaches to flexible working arrangements/remote work are among these factors. Future studies will be needed to understand the impact of these factors on work and health.

## Conclusion

During the COVID-19 pandemic many people either lost their jobs, were temporarily stood-down from work, or furloughed resulting in negative mental health impacts. We have shown that these forms of work cessation contribute to long-lasting poor health outcomes, and that returning to work benefits mental health and may reduce physical activity in the short-term. Provision of timely mental health supports and services for people experiencing work loss will help to reduce the short and potential long-term impacts of work loss on health. Our findings suggest that approaches that minimise the number of workers experiencing work loss will also be beneficial for health. These may include programs that apportion reduced workloads across multiple staff, redeployment of workers into alternative roles, and financial supports that encourage retention of an employment relationship. Services and systems engaging with the newly unemployed should incorporate screening for mental health problems and provide access to mental health services and supports

## Data Availability

The data are held at Monash University, Insurance Work and Health Group, School of Public Health and Preventive Medicine. Procedures to access data from this study are available through contacting the lead author. Proposals for collaborative analyses will be considered by the study's investigator team.

